# The diagnostic accuracy of nucleic acid point-of-care test for human coronavirus: A systematic review and meta-analysis

**DOI:** 10.1101/2020.07.09.20150235

**Authors:** Pakpoom Subsoontorn, Manupat Lohitnavy, Chuenjid Kongkaew

## Abstract

Many recent studies reported coronavirus point of care tests (POCTs) based on isothermal amplification. However, the performances of these tests have not been systematically evaluated. We searched databases for studies that provide data to calculate sensitivity, specificity and diagnostic odds ratio (DOR). We included 43 studies on 5204 specimens. Most studies had high risk of patient selection and index test bias but low risk in other domains. Most studies (n = 21) used reverse transcribed loop mediated isothermal amplification (RT-LAMP) to diagnose Coronavirus disease 2019 (COVID-19). Summary estimated ln(DOR) for RT-LAMP of RNA purified COVID-19 samples is 6.50 (95%CI 5.25-7.76), similar to previously reported value for RT-LAMP of other RNA virus. RT-LAMP from crude samples has significantly lower ln(DOR) at 4.46 (95%CI 3.53-5.38). SAMBA-II has the highest ln(DOR) at 8.00 (95%CI 6.14-9.87). Abbott ID Now performance is similar to RT-LAMP of crude sample. The performances of CRISPR diagnosis and RT-LAMP are not significantly different. Types of coronaviruses and publication status have no significant effect on diagnosis performance. Existing nucleic acid POCTs, particularly RT-LAMP, CRISPR diagnosis and SAMBA-II, have good diagnostic performance. Future work should focus on improving a study design to minimize the risk of biases.

## Introduction

Coronavirus pandemic has caused serious damage to public health and the global economy. Severe acute respiratory syndrome coronavirus (SARS-CoV) and Middle East respiratory syndrome coronavirus (MERS-CoV) infected over ten thousand people and killed over a thousand people worldwide [1]. A novel coronavirus (SARS-CoV-2) that causes coronavirus disease 2019 (COVID-19) infected over 9 million people and killed over 400,000 people (as of June 25th, 2020). The global GDP is predicted to shrink by almost one percent [2]. Rapid and low-cost diagnostic screening of a population at risk is critical for controlling sources of infection. Such diagnostic capability also helps policy makers decide when and to what extent to ease restrictions and restore the economy [3].

Reverse transcribe quantitative polymerase chain reaction (RT-qPCR) has been the gold standard for RNA virus detection [4, 5]. Nonetheless, RT-qPCR requires up to 4 hour sample-to-result time and needs to operate on a bulky expensive thermal cycler with fluorimetry. To fulfill the demand for rapid diagnosis in the outbreak situation, there is a need for point-of-care tests (POCTs) that are cheaper, faster and deployable outside a standard medical laboratory.

Nucleic acid detections based on isothermal amplification obviate the need for a thermal cycler thereby simplifying and speeding up the diagnosis process. For instance, loop-mediated isothermal amplification (LAMP) relies on strand displacing DNA polymerase and primers to amplify specific DNA sequences of pathogens [6]. Reverse transcription LAMP (RT-LAMP) has been applied for the detection of various RNA viruses including Ebola virus, Zika virus, West Nile virus, Influenza virus and Yellow fever virus [7-11]. Rolling circle amplification (RCA) utilizes highly processive strand displacement DNA polymerase and circularizable oligonucleotide probes for detecting single strand DNA or RNA [12]. Reverse transcription insulated isothermal PCR (RT-iiPCR) relies on a temperature gradient to drive denaturation/annealing/extension cycle similar to conventional PCR but in the absence of a thermal cycler [13]. Reverse transcription recombinase polymerase amplification (RT-RPA) or reverse transcription recombinase aided amplification (RT-RAA) uses recombinase, single strand binding protein, DNA polymerase and reverse transcriptase to amplify RNA target [14]. Simple amplification based assay (SAMBA) uses DNA dependent RNA polymerase and RNA dependent DNA polymerase to alternately transcribe and reverse transcribe RNA target [15]. CRISPR diagnosis combines isothermal amplification techniques (such as RT-LAMP) with specific DNA or RNA targeting ability of crRNA and Cas12 or Cas13 enzymes [16]. The outputs of these detection techniques can be coupled with fluorescent or colorimetric reporters as well as lateral flow strip platforms to facilitate readout processes.

While many studies presented nucleic acid POCTs for human coronavirus, it is important to systematically evaluate and draw conclusions about the performance of POCTs and quality of these studies. This could guide clinical practice and highlight opportunities for next generation POCTs. Here, we aim to determine the accuracy of nucleic acid point-of-care diagnosis for human coronavirus, particularly, SARS-CoV, MERS-CoV and SARS-CoV-2, using systematic review and meta-analysis techniques.

## Methods

### Eligibility criteria

This systematic review and meta-analysis included both peer-reviewed and preprint original articles on nucleic acid based POCTs. The test must be isothermal, i.e., thermal cycling is not required during the test. Included studies must have full text available (in any language) and provide enough information to determine the number of true positive, false positive, false negative and true negative on POCTs (performed on clinical samples) relative to a standard reference test.

### Search strategy

Peer-reviewed articles were searched on PubMed from its inception up to 16 June 2020 with the following search terms: (coronavirus OR COVID-19 OR severe acute respiratory syndrome OR middle east respiratory syndrome) AND (rapid diagnosis OR isothermal amplification). Preprint articles were searched on BioRxiv and MedRxiv from 1 January 2020 to 16 June 2020 using a search term ‘isothermal amplification’. The titles and abstracts were screened and the full text of relevant articles were reviewed. We registered our systematic review and meta-analysis on PROSPERO on April 21, 2020; registration number to be updated.

### Quality assessment

The quality of each study was assessed with the Quality Assessment of Diagnostic Accuracy Studies 2 (QUADAS-2) [17]. QUADAS-2 determines the risk of bias and the applicability of each study in four main areas: patient selection, index test, reference standard, and flow and timing. These domains were assessed by using 18 signaling questions with yes, no and unclear answers. Then, the answers were used to judge whether the risk of bias and the concern for the applicability of the research is low, high or unclear. Two reviewers (PS and CK) independently judged the quality of each study. Disagreements were resolved by consensus with additional input from the third party (ML).

### Data extraction

Data were extracted by one reviewer (PS) and where the results were unclear the two other reviewers (CK and ML) were consulted. The parameters extracted include: citation information, types of coronavirus, methodology, and the diagnostic accuracy results.

### Statistical data analysis and reporting

Heterogeneity between included studies was assessed using c^2^ and *I*^2^ tests to determine whether it was appropriate to compute an estimate of the meta-analytic summary [18-19]. The summary weighted mean differences and 95% confidence intervals (95%CIs) were calculated based on the DerSimonian and Laird method under a random effect model [20]. P-values::0.05 indicated heterogeneity between studies. *I*^2^ values of 25%, 50%, 75% denote a low, moderate, and high degree of heterogeneity across studies. All statistical analyses were performed using the R program (version 3.4.0) [21].

Forest plots were generated using R ‘mada’ [22] and ‘forestplot’ package [23]. To avoid statistical artefacts from having zero cells in a 2×2 table (for example when false positive or false negative are zero), continuity corrections = 0.5 were added to the observed frequencies when calculating diagnostic odds ratio (DOR) [22]. Since pooling sensitivity or specificity can be misleading, only univariate meta-analysis of DOR was calculated using the ‘madauni’ command in ‘mada’ package [24-25].

## Results

### Search results

We identified 1308 articles in total through database searching (Fig 1). After title and abstract screening, we excluded 1225 articles that were not primary research articles, had no full text available or were unrelated to nucleic acid POCTs for human coronavirus. 62 non-English articles were found but only two met above eligibility criteria. These two articles were later excluded as the authors did not use clinical samples. After reviewing full text, we found only 43 articles with sufficient information to calculate sensitivity, specificity and diagnostic odds ratio (DOR) on clinical samples [26-68] (Table 1, Table S1). Among 43 included articles, there were 38 articles on the diagnosis of COVID19, three articles on the diagnosis of MERS and two articles on the diagnosis of SARS.

**Table 1:**
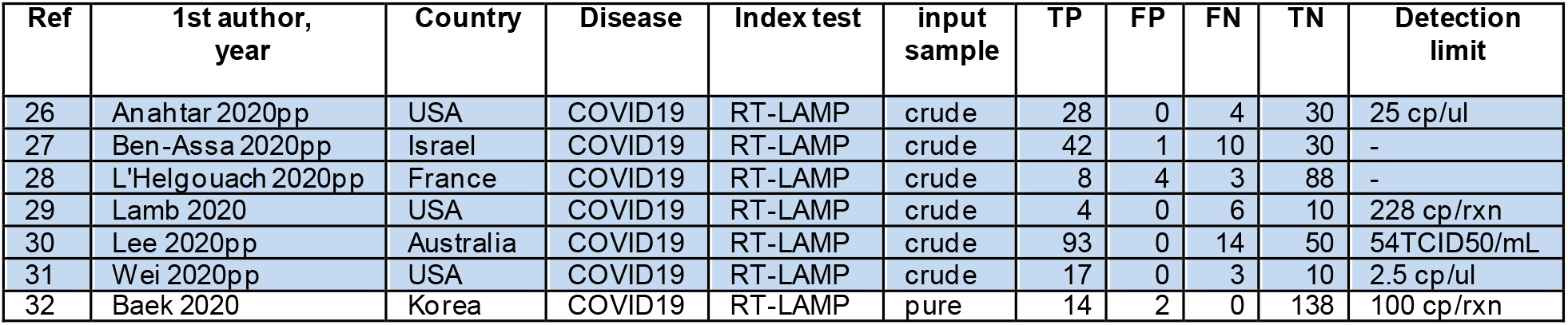
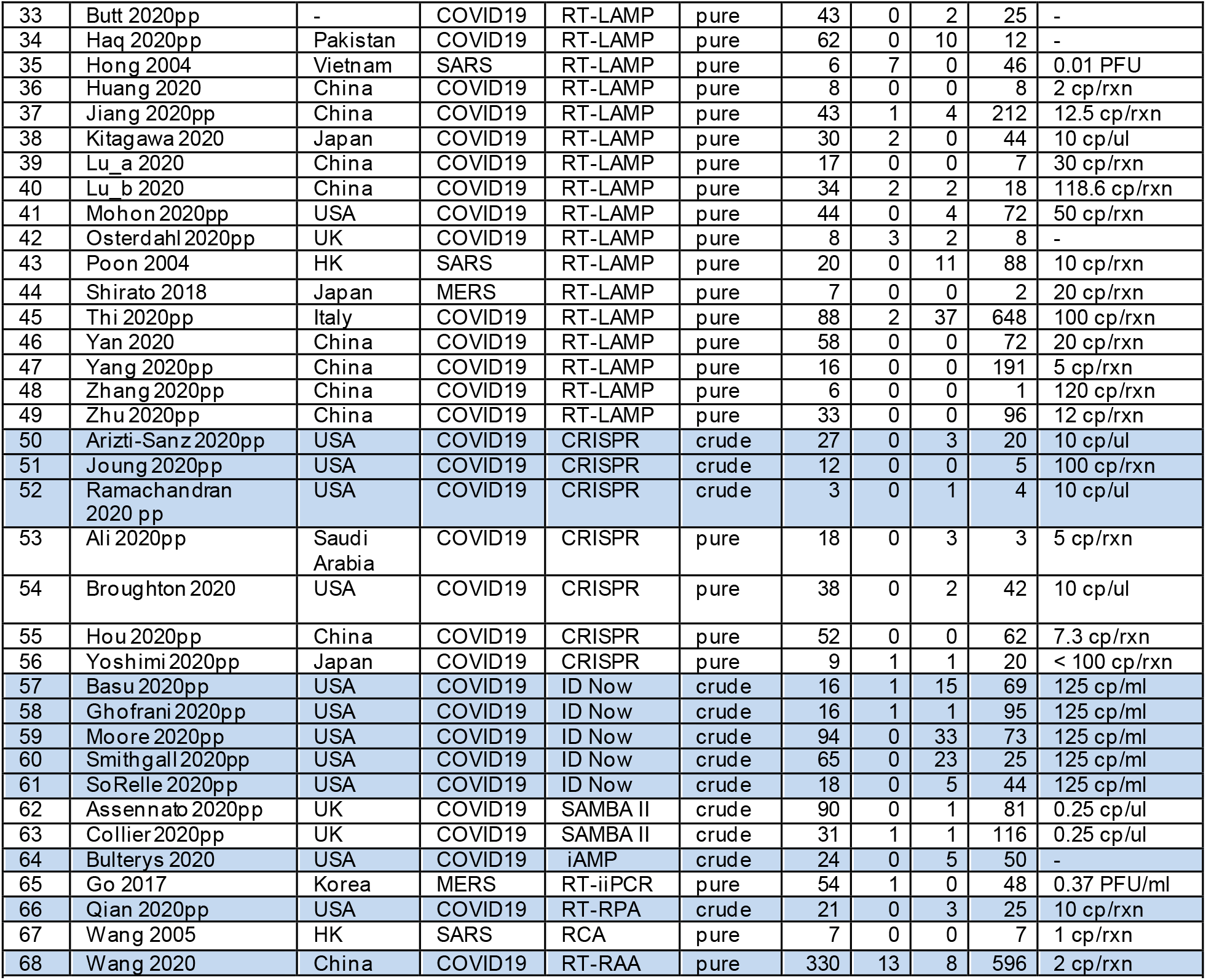
summary of all included studies. Each row for each study. Adjacent rows are shaded differently if the studies shown in those rows used differnt index test or input samples. **Abbreviation:** USA, United State of America; UK, United Kingdoms; HK, Hong Kong. PFU, plaque forming unit; TCID50, 50% tissue culture infective dose; cp/rxn, copies/reaction; TP, true positive; FP, false positive; FN, false negative; TN, true negative. “Pure” indicates that the studies used purified RNA extracted fro m patient samples; “Crude” indicates that the study used crude patient samples. “-” indicates that data was not reported. All studies used RT-qPCR as a reference standard test except for Poon et al 2004 using immunofluorescent assay (IFA) [43] and Hong et al 2004 [35] using non quantitative RT-PCR (readout result in agarose gel). “pp” in the 1st author-year column indicates that the study is in preprint (not peer-reviewed).

**Figure 1.**
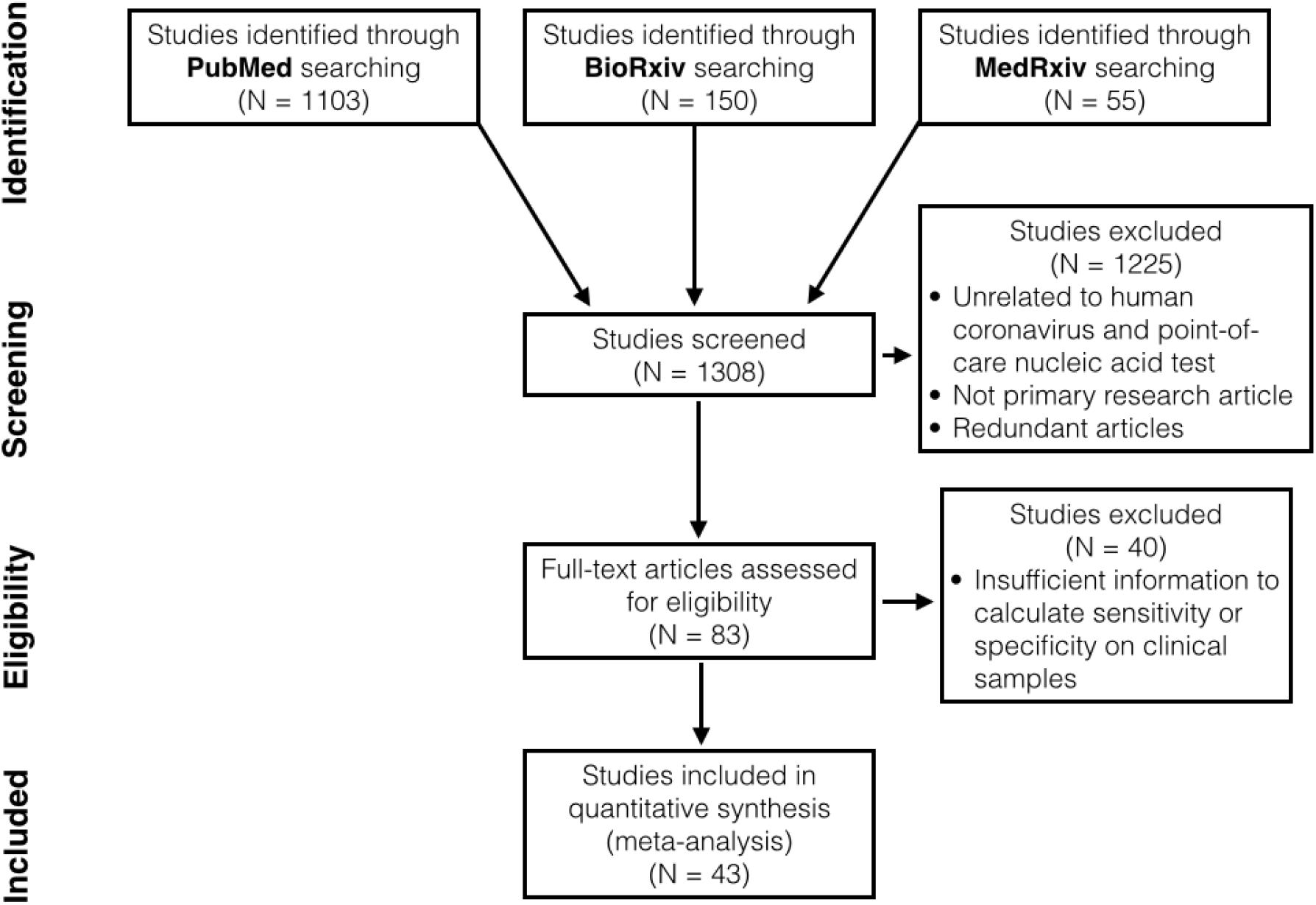
The PRISMA flow diagram.

### Characteristics of the included studies

In total, there were 5204 clinical samples analyzed. Most studies used clinical samples from USA (n = 15 out of 43 studies), followed by China (n = 10), Japan (n = 3), UK (n = 3), Korea (n = 2) and Hong Kong (n = 2). The rest were from Australia, France, Israel, Italy, Pakistan, Saudi Arabia, Vietnam, and an unspecified country (n = 1 each). Most articles (n = 38) were COVID-19 diagnosis studies published or uploaded to preprint databases in 2020. Most studies (n = 24) use RT-LAMP as nucleic acid POCTs, followed by CRISPR diagnosis (n = 7), Abbott ID Now (n = 5) and SAMBA II (n = 2). The rest were iAMP, RT-iiPCR, RT-RPA, RT-RAA and RCA (n = 1 each). Over a third (n = 18) of all studies attempted to diagnose coronavirus in crude patient samples, i.e, nasopharyngeal swabs, sputum, saliva, etc.; the rest used purified RNA from patient samples for diagnosis.

### Quality of articles

Over three fourths of all studies (n = 33 out of 43 studies) have high risk of patient selection bias due to non-random patient selection and case-control study design (Fig 2, Table S2-S3). These studies specifically recruited clinical samples known to be uninfected or infected with coronavirus. Other studies [38, 39, 45, 46, 57, 58, 61, 63, 68] have unclear risk of patient selection bias because these studies were not case-control but provided insufficient detail about patient inclusion/exclusion criteria. Only one study [62] has low risk of patient selection bias.

**Figure 2.**
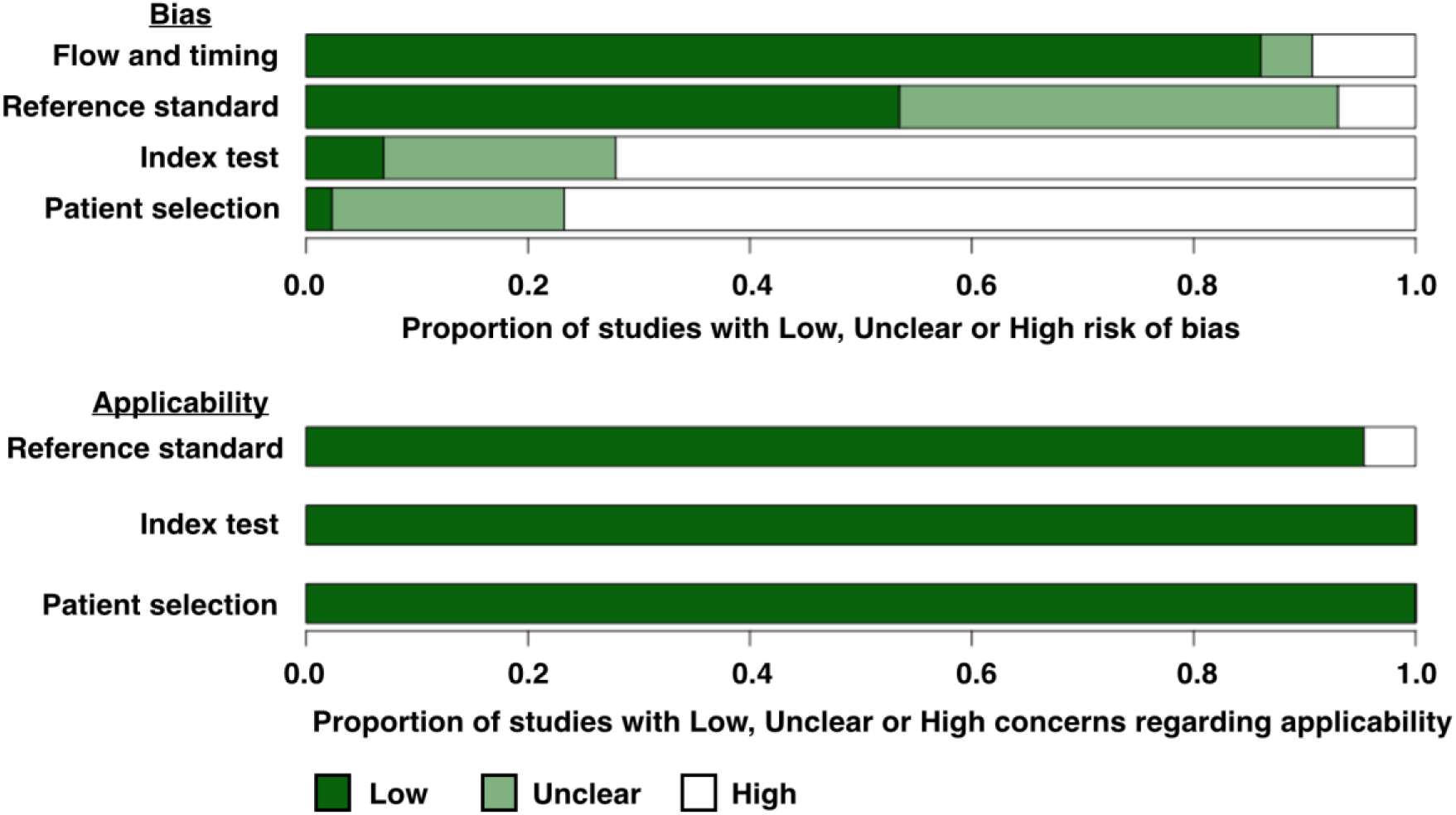
QUADAS-2 finding per domain for 43 studies included in this systematic review.

Almost three fourth of all studies (n = 31 out of 43 studies) have a high risk of index test bias because the index test results were interpreted with knowledge of reference standard results or qualitative readout was used for interpreting the result. For the rest, three studies [37, 46, 68] explicitly stated that index and reference test were done simultaneously/in parallel while nine other studies (∼low risk) [28, 38, 44, 45, 58, 59, 61, 62, 63] did not provide enough information (∼unclear risk).

Only three studies have high risk of reference standard bias. One of these three studies did not use the same RT-qPCR kits and protocols for different patient group [68]. Other two studies used RT-PCR (not quantitative, readout result in agarose gel electrophoresis) [35] or immunofluorescent assay (IFA) [43] as a reference standard test. For the rest of included studies (n = 40), almost half (n = 17) have unclear risk of reference standard bias because these studies did not provide enough information whether reference standard results were interpreted without knowledge of the results of the index test.

Most studies (n = 38 out of 43) have a low risk of flow and timing bias with the following exceptions. One study provided no information on whether the samples for a reference test (IFA) and the index test (RT-LAMP) were taken at the same time [43]. Another study might excluded some samples from the workflow [48]. These two studies were marked as having unknown risk of flow and timing bias. Three studies were designated as having high risk due to the use of different standard references on different samples [68], the use of different samples test flow on different sample groups [42] and the exclusion of some samples from the analysis [44].

Our review question does not focus on any particular patient demographics. None of the 43 included studies attempted to exclude patients based on demographics and thus has no concern of patient selection applicability. Index isothermal tests of all studies have generally been used for POCTs and thus have low concern of index test applicability. Reference standard tests of nearly all studies are RT-qPCR, a gold standard for RNA virus detection. Thus, we graded these studies as having low concern of standard test applicability. Two studies that used (non-quantitative) RT-PCR [35] and IFA [43], were marked as having high concern of standard test applicability.

### Sensitivity and specificity of nucleic acid POCTs

Across all included studies, the sensitivity ranged from 40% to 100%, and the specificity ranged from 73% to 100% (Fig 3A). Almost a quarter of all studies (n = 10 out of 43) reported both 100% sensitivity and 100% specificity. However, most of these ten studies have wide 95% CIs due to small numbers of clinical samples. For instance, the study by Zhang et al (2020) [48] with only seven clinical samples has 95%CIs of sensitivity and specificity at 61-100% and 21-100%, respectively. Only two studies by Hou et al and Yan et al reported 100% sensitivity and specificity with 95%CIs less than 10% [55, 46]. Yan et al demonstrated 100% consistency between RT-LAMP and RT-qPCR test results on 130 clinical samples. Notably, this study is also one of the highest quality studies in our review, having unknown risk of bias in only one QUADAS-2 domain and low risk of bias or applicability in all other domains. Hou et al reported 100% consistency between CRISPR diagnosis system and RT-qPCR test results on 114 clinical samples. However, unlike the study by Yan et al, this study did not use random sample selection and the index (CRISPR diagnosis) test results were interpreted with the knowledge of reference standard. In other words, Hou et al study has higher risk of bias than Yan et al study.

**Figure 3.**
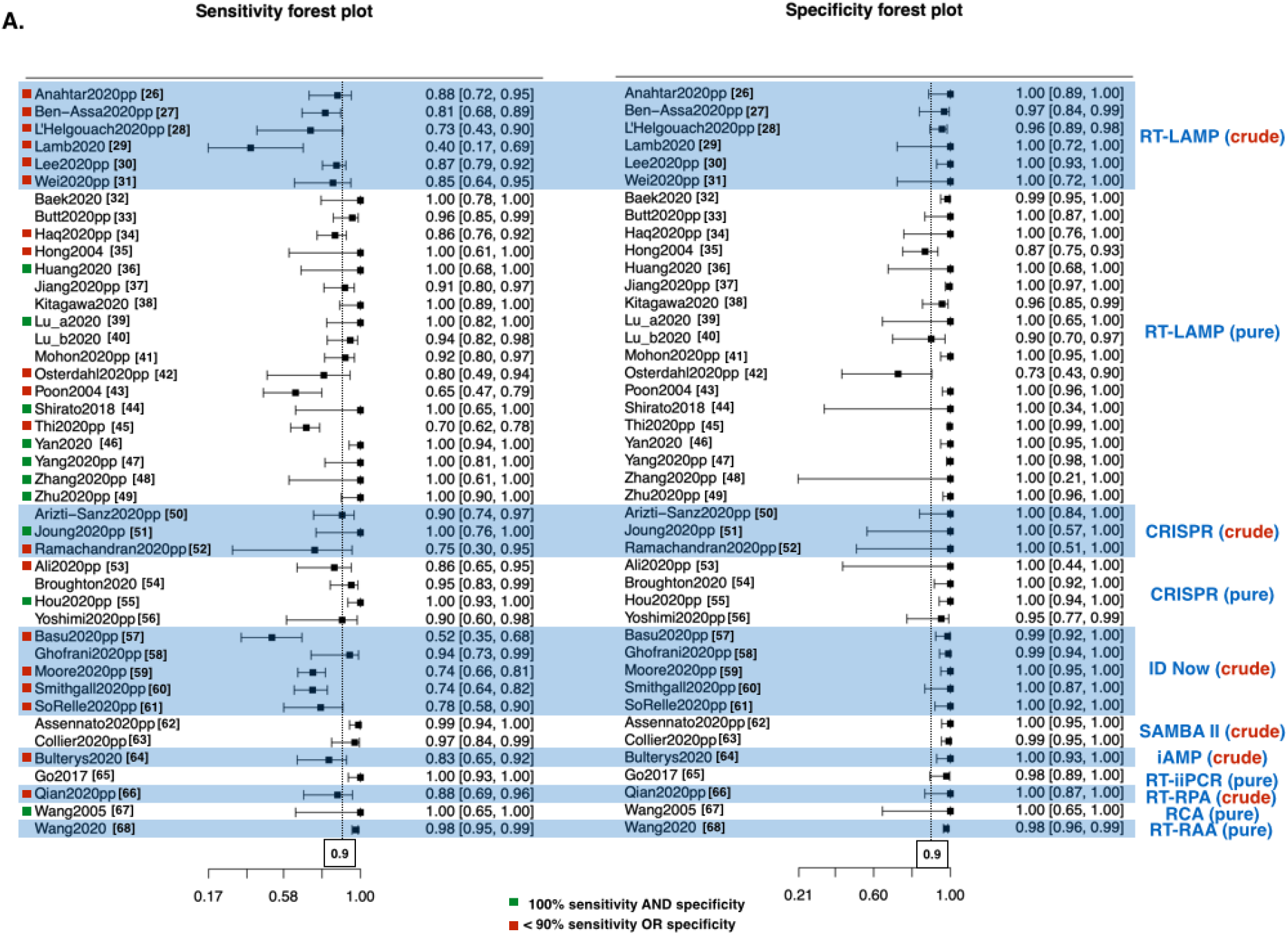
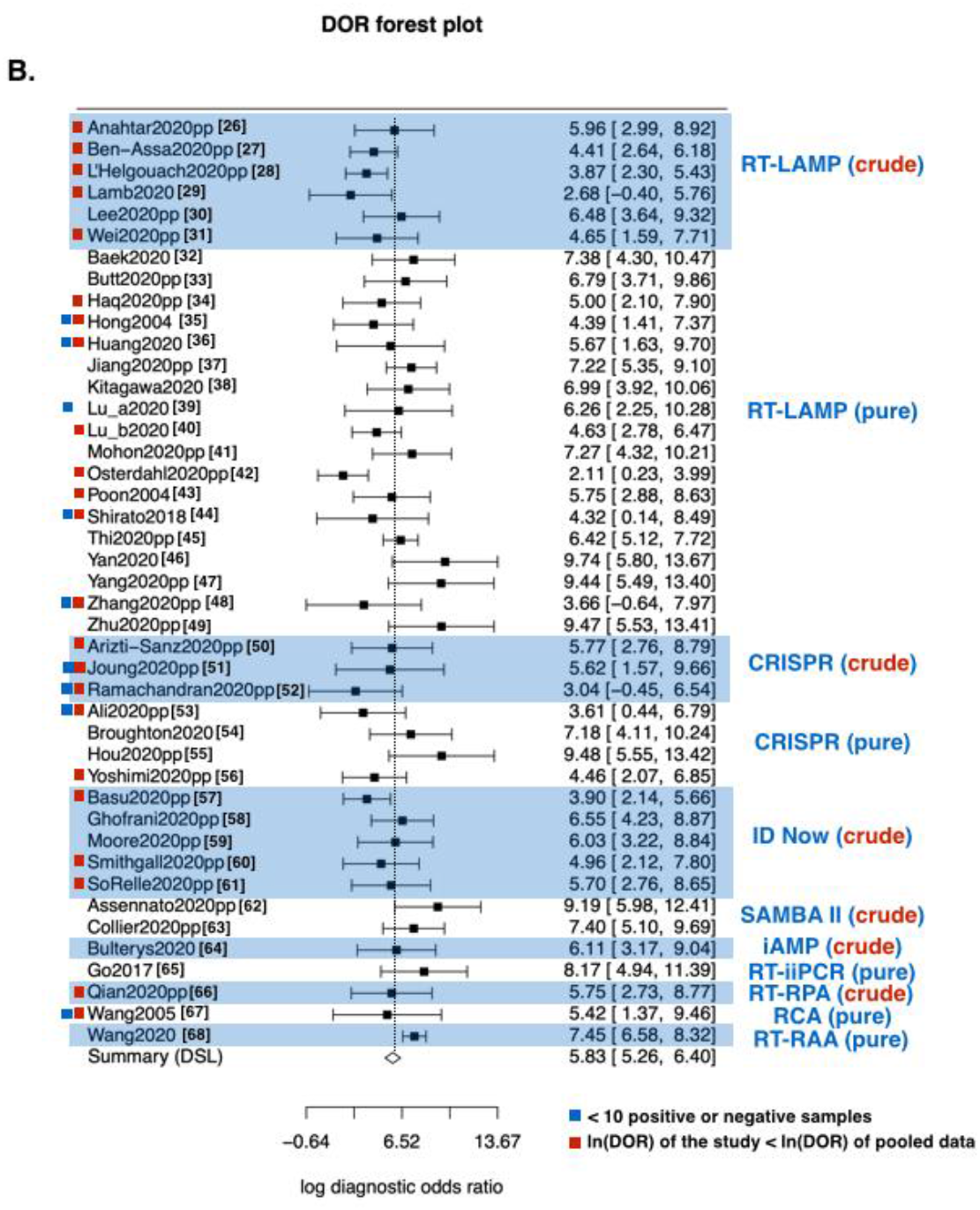
The forest plot of sensitivity, specificity and diagnostic odds ratio (DOR) of human coronavirus nucleic acid POCTs. **(A) Sensitivity and Specificity.** Each row shows the first author name and publication year of each study. Behind publication year, “pp” indicates that the study is preprint (not peer-reviewed). Vertical dotted lines indicate 90% sensitivity or specificity. **(B) Diagnostic Odds Ratio (DOR)**. The vertical dotted line denotes ln(DOR) of pooled data from all studies

Over a third of the included studies (n = 19 out of 43) reported less than 90% sensitivity while only two studies reported less than 90% specificity. Over two third (n = 13 out of 18) of studies that used crude patient samples for diagnosis reported less than 90% sensitivity. On the contrary, only a fifth (n = 5 out of 25) of studies that used purified RNA reported less than 90% sensitivity. Among the six studies that used purified RNA samples but has less than 90% sensitivity or specificity [34, 35, 42, 43, 45, 53], three studies have unusual QUADAS-2 risks and concerns, comparing other included studies. Hong et al [35] and Poon et al [43] had high risk of reference standard bias and concern of standard applicability. This is because these two studies used RT-PCR and IFA instead of RT-qPCR for reference standard. Osterdahl et al [42] has only 20 clinical samples. Moreover, the study by Osterdahl et al is one of the only three studies marked as having high risk of flow and timing bias because some clinical samples were taken on different days for index test and standard reference test. Notably, the study that reported the lowest sensitivity among those that used purified RNA samples (at 70%) is also one of the best quality study: no high risk/concern in any QUADAS-2 domain and having more tested samples than any other study in this group [45].

### Meta-analysis of diagnostic odds ratio (DOR)

The included studies have ln(DOR) ranging from 2.11 to 9.74 (Fig 3B). Pooled data from all included studies (n = 43) has ln(DOR) of 5.83 (95% CI 5.26 - 6.40) with no observed heterogeneity (X^2^ =37.71, d.f.=42, p=0.657; *I*^*2*^ =0%). The top two performers [46, 55] with regard to sensitivity and specificity also have highest ln(DOR). About half of all studies (n = 23 out of 43) have ln(DOR) below that of the pooled data. Eighteen of these studies are those with sensitivity or specificity no more than 90% [26-29, 31, 34, 35, 40, 42, 43, 50, 52, 53, 56, 57, 60, 61, 66]. The other five studies have low ln(DOR) despite having 100% sensitivity and 100% specificity [36, 44, 48, 51, 67]. These studies have small numbers of test samples so continuity correction during DOR calculation significantly reduced ln(DOR).

Subgroup analyses were performed based on methods of detection, types of diseases, numbers of tested samples and publication statuses (Fig 4). All analyzed subgroups have ln(DOR) above zero; almost all analyzed subgroups have no heterogeneity (*I*^*2*^ =0%) except CRISPR diagnosis of purified sample (*I*^*2*^ =9.522%, low heterogeneity). Among different methods of detection with multiple studies, SAMBA II has the highest ln(DOR) at 8.00 followed by RT-LAMP and CRISPR diagnosis of purified samples (ln(DOR) at 6.06 and 5.94, respectively).

**Figure 4.**
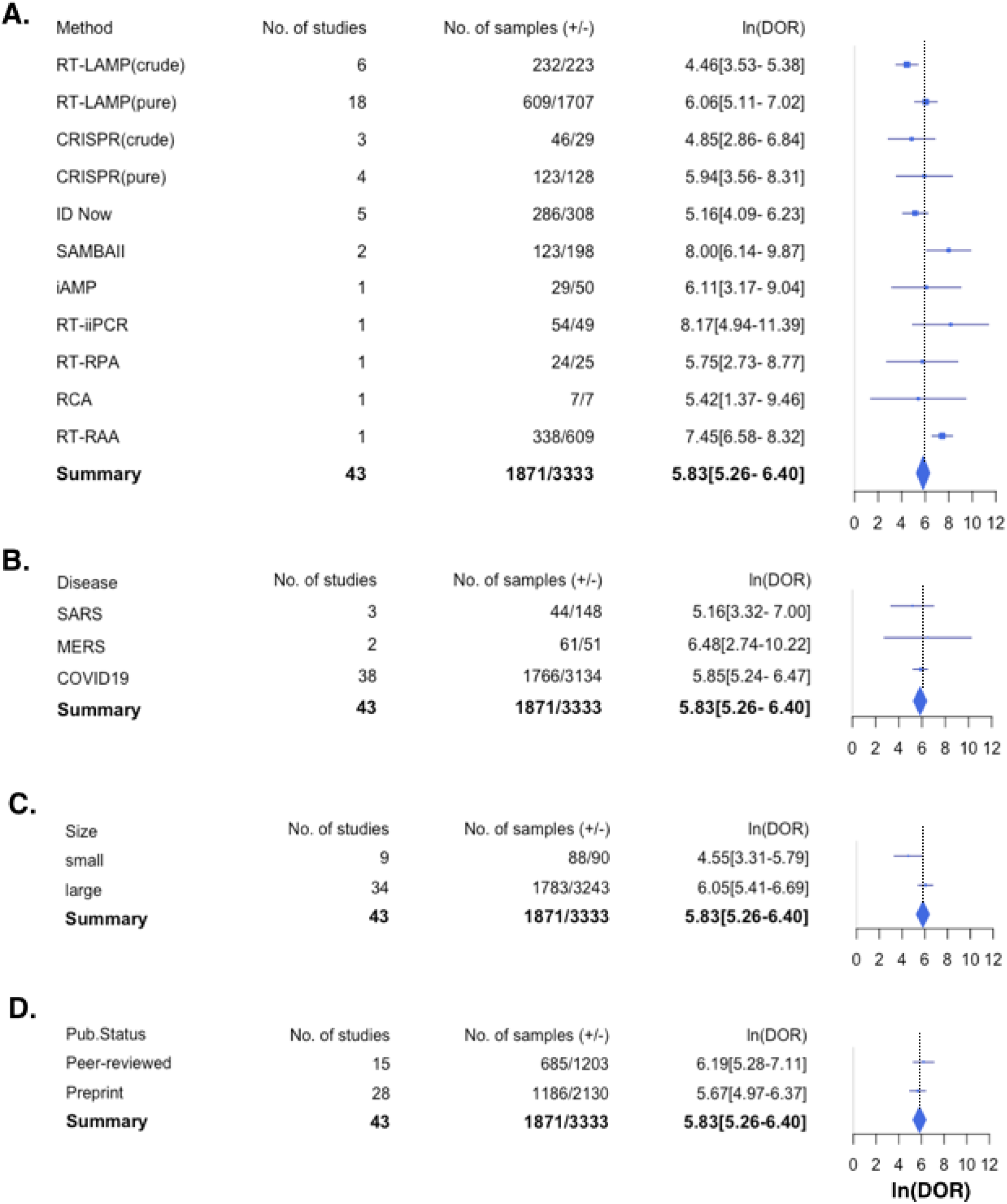
Subgroup analyses of all included studies. **S**tudies were grouped by **(A) v**iral RNA detection methods, **(B)** types of coronavirus diseases (SARS, MERS and COVID-19), **(C)** specimen numbers (less than 10 or at least 10) and **(D)** publication status (peer-reviewed and preprint). A study is in the “less than 10” group if it has less than 10 human coronavirus infected clinical samples OR less than 10 uninfected samples. A study is in the “at least 10” group if it has at least 10 coronavirus infected clinical samples AND at least 10 uninfected samples.

RT-LAMP of crude samples have the lowest ln(DOR) at 4.46, followed by CRISPR diagnosis of crude samples and ID Now (ln(DOR) at 4.85 and 5.16, respectively). Comparing among different diseases, MERS diagnosis has the highest ln(DOR) at 6.48, followed by COVID19 diagnosis and SARS diagnosis (ln(DOR) at 5.85 and 5.16, respective). Studies with large numbers of test samples (at least 10 positive and negative samples) has higher ln(DOR) than studies with smaller numbers of test samples (less than 10 positive or negative samples) (ln(DOR) at 6.05 and 4.55, respectively). Note that this difference is likely to result from the fact that continuity correct has more impact on studies with smaller sample sizes. Preprint studies have less ln(DOR) than peer-reviewed studies (ln(DOR) = 5.67 and 6.19, respectively).

For subgroup analyses based on types of diseases and publication status, differences in ln(DOR) among compared subgroups are small, i.e., within 95% CIs of one another. For example, ln(DOR) of peer-review subgroup (6.19) is within 95% CI of ln(DOR) of preprint subgroup (4.97-6.37). On the hand, ln(DOR) of preprint subgroup (5.67) is within 95% CI of ln(DOR) of peer-reviewed subgroup (5.28-7.11). For subgroup analysis based on diagnosis methods, ln(DOR) of RT-LAMP and CRISPR diagnosis are within 95% CI of each other. However, ln(DOR) of RT-LAMP of purified samples and of crude samples are not within 95% CI of each other. ln(DOR) of ID Now is barely within 95% of RT-LAMP of purified RNA samples while ln(DOR) of SAMBA II is well above 95% CI of purified RNA samples.

We performed further subgroup analyses using only data from COVID-19 diagnosis studies with at least 10 samples for infected and uninfected sample groups (Fig 5). All analyzed subgroups have ln(DOR) above zero; almost all analyzed subgroups have no heterogeneity (*I*^*2*^ =0%). Among different methods of detection with multiple studies, RT-LAMP of crude sample still have the lowest ln(DOR) at 4.62 followed by ID Now at 5.16 (both ln(DOR) are within 95% CI of each other). SAMBA II still has the highest ln(DOR) at 8.00. Nonetheless, ln(DOR) of SAMBA II, RT-LAMP of purified RNA and CRISPR diagnosis of purified or crude RNA are within 95% CI of one another. Note that there is only one CRISPR diagnosis study on crude samples left in this analysis. Within the study group using RT-LAMP of purified RNA, we further analyzed the difference between peer-reviewed and preprint studies (Fig 5). The peer-reviewed and preprint groups have almost equal ln(DOR) at 6.77 and 6.41; these ln(DOR) values are within 95% CI of each other.

**Figure 5.**
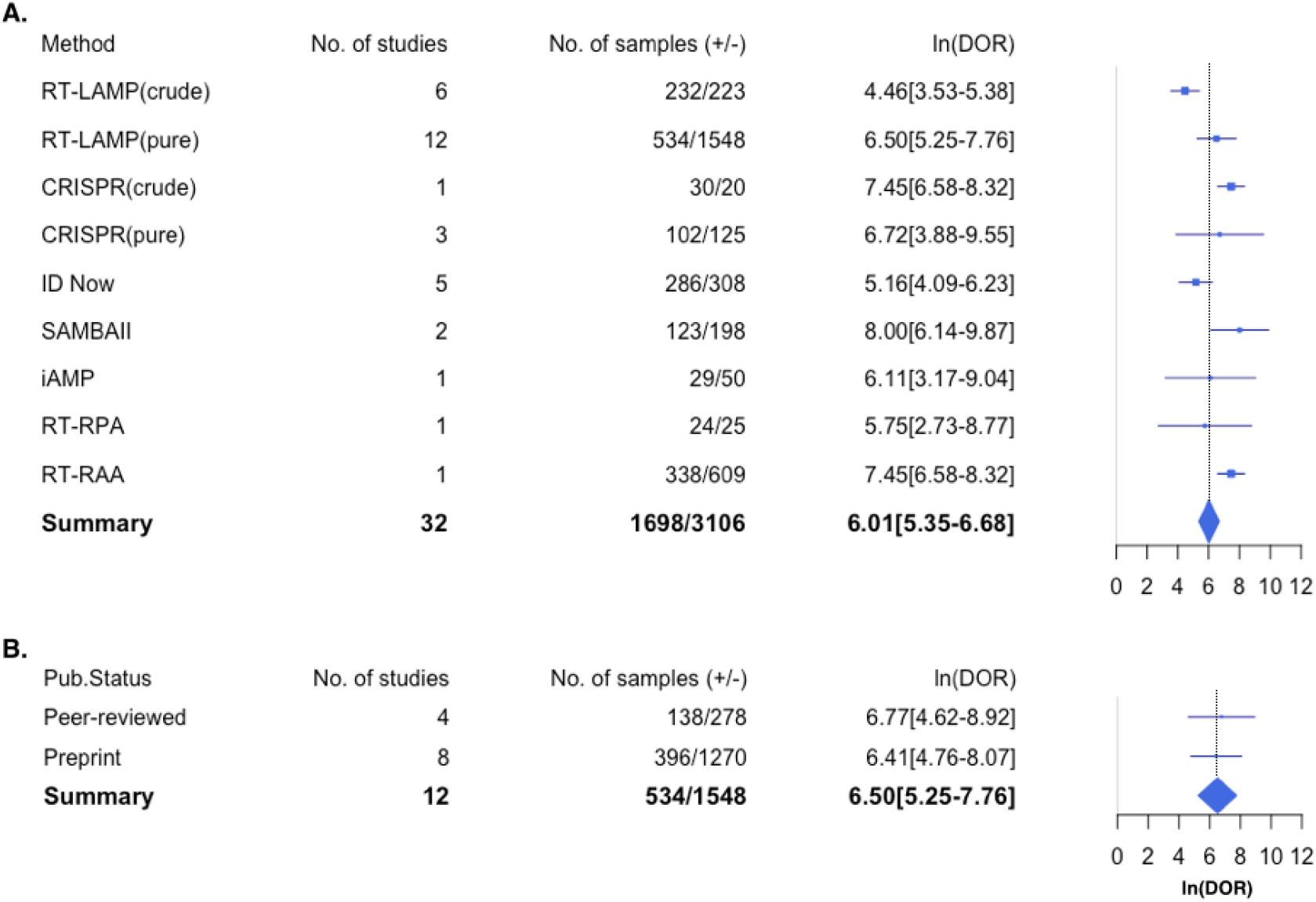
Subgroup analyses of COVID-19 diagnosis studies with large numbers of tested samples. All studies in this analysis must also have at least 10 coronavirus infected clinical samples AND at least 10 uninfected samples. (A) subgroup analysis based on diagnosis methods, **(B)** subgroup analysis based on publication status of RT-LAMP studies on purified RNA samples.

## Discussion

This is the first systematic review and meta-analysis examining the performance of nucleic acid POCTs for human coronavirus. Out of 43 studies with 5204 patient samples in total, over half (n = 24) of all included studies reported over 90% sensitivity and specificity; almost a quarter (n = 10) of all included studies reporter 100% sensitivity and specificity. ln(DOR) of each and pooled study data are high indicating overall good diagnosis accuracy. Subgroup analyses showed that the type of coronavirus and the publication status has no significant impact on ln(DOR). Studies with smaller number of samples have significantly lower ln(DOR) than studies with larger number of samples. However, six out of nine studies in the small samples size subgroup reported 100% sensitivity and specificity. Thus, the observed difference ln(DOR) between subgroup with small sample number and large sample number is likely to result from continuity correction rather than diagnostic performance of the test itself. Based on data from our included studies, the key determinants of diagnosis accuracy is the type of assay and whether crude sample or purified RNA was used for diagnosis.

At the time of this writing, the only published meta-analysis study on the accuracy of isothermal nucleic acid test for RNA virus is on RT-LAMP performance for Enterovirus 71 [69]. The meta-analysis includes 907 clinical samples from 10 studies, all performed on purified RNA. Pooled data have ln(DOR) of 6.74 (95% CIs 5.68-7.79) with no observed heterogeneity. This level of performance for Enterovirus detection was almost equal to our calculated ln(DOR) of RT-LAMP for purified RNA from COVID-19 patients (ln(DOR) = 6.50, 95% CI of 5.25-7.76, I^2^ = 0%). Thus, our reported RT-LAMP performance is likely to reflect the true performance of this isothermal nucleic acid test as the performance value is generalisable across different target viruses. This could serve as a reference point for assessing the performance of other diagnosis methods.

Among RT-LAMP studies on purified RNA, the studies by Kitakawa et al., Thi et al. and Yan et al [38, 45, 46] are of the highest quality: large number of tested samples and no QUADAS-2 domain with high risk of bias or concern of applicability. Notably these three studies reported contrasting results with respect to diagnosis performance. While Kitakawa et al [38] and Yan et al [46] demonstrated 100% diagnostic sensitivity, Thi et al [45] reported only 70% sensitivity. Generally, the sensitivity of diagnosis test decreases when concentration of viral RNA in the samples decreases. For example, Thi et al showed that RT-LAMP sensitivity is at 100% when the samples have viral RNA concentration equivalent to Ct ∼ 0-25. The sensitivity decreases to about 30% at RNA concentration Ct∼30-35 and to sensitivity less than 6% at RNA concentration ∼ 35-40. Approximately a third of positive samples in Thi et al study has Ct ∼ 30-This could explain why RT-LAMP in this study appear to have such a low overall sensitivity. Yan et al and Kitakawa et al did not report the distribution of viral RNA level in their tested samples. Thus, it is possible these two studies appear to achieve 100% sensitivity simply because most of their positive samples had high viral RNA level.

Viral RNA levels in samples depend on several factors including severity of the disease, sample collection timing, type of samples and sample handling process. Without such information, it is difficult to determine whether the difference in observed sensitivity results from the performance of the test itself or the properties of the samples used in the test. Unfortunately, most included studies provided no information about viral RNA level in the infected samples (as determined by a standard reference test, e.g., RT-qPCR). Information about disease severity and sample collection timing (i.e. days after disease onset) are often missing. Future work should provide this information in order to allow better assessment of diagnosis test performance and identify their actual limitations.

No published study thus far directly compared the Cororavirus detection accuracy of CRISPR diagnosis to that of RT-LAMP. Subgroup analysis showed that, for purified RNA samples, ln(DOR) of CRISPR diagnosis is only slightly higher than that of RT-LAMP (ln(DOR) within 95% CI of each other) (Fig 4A, 5A). Additionally, reported limits of detection of these CRISPR diagnosis tests (7.3-100 copies/reaction) are within the same range as that of RT-LAMP (2-120 copies/reaction) (Table 1). Given available data, it cannot be concluded that CRISPR diagnosis can outperform RT-LAMP at least for purified RNA samples. Existing CRISPR diagnosis also requires RT-LAMP or other isothermal techniques to pre-amplify nucleic acid targets before CRISPR detection. The use of cas12 or cas13 enzyme adds to the cost of CRISPR diagnosis test kit, making it likely to be more expensive than RT-LAMP. Future study should directly compare and highlight unique strength of CRISPR diagnosis relative other isothermal techniques, for example, its ability for multiplex detection and identifying single base difference in targeted genomes [70-72].

Our systematic review and meta-analysis also included one study each on diagnosis accuracy of RT-iiPCR, RCA and RT-RAA using purified RNA samples. The two studies using RCA and RT-iiPCR reported perfect or near perfect sensitivity and specificity [65, 67]. However, the number of tested samples in these studies was too small to draw conclusion about their diagnosis performance relative to other POCTs. On the contrary, the study using RT-RAA had almost a thousand tested samples, the largest number among all 43 studies in our systematic review [68]. This study was peer-reviewed and has only one high risk of bias in reference standard domain as different standard tests were applied to different patient groups. Still, these reference standard tests were merely different variants of RT-qPCR. This study reported up to 98% sensitivity and specificity and had the narrowest 95% CI among all included studies. Subgroup analysis implies that RT-RAA accuracy is significantly higher than that of RT-LAMP on purified samples (CI 95% confident interval not overlapping each other). Future work should directly compare this assay to other nucleic acid POCTs such as RT-LAMP using the same sample set in order to determine the actual difference in diagnosis performance.

Nearly all published peer-reviewed studies (13 out of 15 studies) applied diagnostic assays on viral RNA purified from patient samples. However, RNA extraction step increases assay cost and time and often requires trained personnel. In order to accommodate large scale field deployable POCT, more recent studies aimed to develop assays that can bypass or automate this purification step. Our meta-analysis showed that diagnosis accuracy of patient samples is still generally lower than that of purified RNA (Fig 4A and 5A). All six studies that used RT-LAMP on crude patient samples report sensitivity or specificity at 90% or below. On the contrary, the majority (14 out of 18) studies that used RT-LAMP on purified RNA samples reported over 90% sensitivity and specificity; half of these studies reported 100% sensitivity and specificity. Pooled data from studies using RT-LAMP on purified RNA also have higher ln(DOR) than that from studies using RT-LAMP on crude samples. Additionally, studies on crude sample diagnosis with other methods including iAMP, RT-RPA, and ID Now reported low sensitivity, specificity and pooled ln(DOR) (Fig 3A). For CRISPR diagnosis, our include studies showed that the diagnosis accuracy of crude samples did not significantly differ from that of purified RNA (Fig 4A and 5A). One studies even reported 100% sensitivity and specificity of CRISPR diagnosis on crude sample. Nonetheless, the number of studies and of tested samples is still too small to drawn solid conclusion on whether accurate diagnosis on crude sample has already been achieved.

Other studies on crude patient samples uses RT-RPA [66], ID Now [57-61] and SAMBA II [62-63] as diagnosis methods. The study using RT-RPA reported ln(DOR) value between that of RT-LAMP crude sample and RT-LAMP purified RNA. However, we found only one RT-RPA study for this review and the sample size of this study is quite small (< 50 total samples). Thus, we cannot draw conclusion about the performance of this methods relative to other methods. Abbott ID Now is famous for being “the fastest” (5-13 minute) isothermal COVID-19 nucleic acid detection system in the market. However, four out of five ID Now studies included in our review reported less than 80% sensitivity. Pooled data from ID Now studies have ln(DOR) level on par with ln(DOR) of RT-LAMP applied to crude samples. The two SAMBA II studies each includes over a hundred tested samples and have no high risk or concern QUADAS-2 domains [62-63]. Each of these two studies sensitivity and specificity at 97% or above while their pooled data has the highest ln(DOR) among all diagnostic assays in our review (including those that use purified RNA). Despite being the slowest POCTs among our included studies (> 1 hr from sample to readout), SAMBA II is arguably the most promising POCTs thus far regarding diagnosis accuracy of coronavirus detection.

Our study identifies both relevant peer-reviewed studies and preprints for deriving better scientific conclusions in diagnosis of the life-threatening novel coronavirus in a timely manner. Our study also adheres to standard methodology of systematic review and meta-analysis as indicated by the PRISMA statement [73]. However, our study has a few limitations. First, most of the included studies (n=29 out for 43) have high risk of patient selection bias and index test bias. Such bias could lead to overestimation of diagnosis performance. Nonetheless, the study that reported the highest performance (near 100% sensitivity and 100% specificity with narrow 95% CIs) was also the one with lowest QUADAS risk and concerns in all domains [38, 46, 62, 63, 68]. Second, almost a forth (n = 9) of included studies have small numbers of test samples. Consequently, these studies have wide 95% CIs of sensitivity and specificity and have DOR highly distorted by continuity correction. Nonetheless, our main conclusion about relative performances for difference diagnosis methods does not change when these studies were excluded. Third, almost two third (n = 28) of included studies have not been peer-reviewed. Nevertheless, our analysis shows that DOR for pooled data from these preprint manuscripts is not significantly different from that of published manuscripts. Therefore, an inclusion of data from preprint manuscripts is unlikely to skew the results of our other analysis. Given that the peer-review process often takes at least a few months, the systematic review that includes preprint manuscripts could be necessary for guiding the direction of on-going research especially during a global pandemic.

In conclusion, our systematic review and meta-analysis reveals the current state of nucleic acid POCTs for human coronavirus. Overall diagnosis accuracy of these POCTs reported so far is high but the quality of these studies was still in question. Despite high diversity of detection methods and RNA target, heterogeneity across studies are low. There is no significant difference in diagnosis performance for different coronaviruses and whether the studies have been published or still in preprint stage. Critical information about viral load or factors influencing viral load was missing in most studies. It is still unclear whether CRISPR diagnosis is superior to a cheaper, simpler and more established nucleic acid POCTs such as RT-LAMP. SAMBA II has highest diagnostic accuracy among all POCTs in this systematic review while Abbott ID Now has lower diagnostic accuracy The performance of viral detection directly from patient samples is significantly lower than from purified RNA. The success in bypassing this RNA extraction step will simplify the workflow, reduce time, cost and possible error. The improvement in these key areas will bring nucleic acid POCTs toward large practical uses for surveillance of on-going and future coronavirus outbreaks.

## Data Availability

All relevant data are in the manuscript or supplementary materials.

## Supporting Information

See supplementary table S1, S2, S3

